# Determinants of Under-nutrition among Children Aged 6 up to 59 Months in Akaki Kaliti Sub City, Addis Ababa: Unmatched Case-Control Study

**DOI:** 10.1101/2023.01.25.23285000

**Authors:** Abeje Kebede, Dagmawit Getaneh, Derbachew Asfaw, Addis Birhanu

## Abstract

**Background:** Malnutrition remains a major health problem and globally it is responsible for one-third of all infant and child mortality. Despite progress has been made in improving child nutrition in Ethiopia in the last decades, it remains a problem of public health importance. Though, there were numerous studies undertaken previously, no studies were previously tried to verify the determinants of malnutrition among children age 6 to 59 months in the study area. Hence, there is a necessity to carry out a study to come up with the determinant factors of malnutrition in the Akaki Kaliti sub-city, Addis Ababa.

**Objective of the Study:** To identify determinants of undernutrition among children age 6 to 59 months in Akaki Kaliti Sub-city, Addis Ababa, 2021.

**Methods and Materials:** Facility-based unmatched case control study was conducted in public health facilities of Akaki Kaliti Sub-city, Addis Ababa from April 26 to May 31, 2021. Consecutive sampling technique was employed. Primary data was collected using interviewer-administered structured questionnaire and analyzed using SPSS for windows version 21. Variables with P-value <0.25 in the bivariate analysis were identified and fitted in multivariable logistic regression model to identify independent predictors of child undernutrition. The magnitude of association was measured by AOR with 95% confidence interval. A P-value less than 0.05 were used to declare the observed association is statistically significant.

**Result:** A total of 83 children (cases) and 159 children (controls) with their mothers/care givers were participated in the study. Having upper respiratory tract infection in the past two weeks [AOR = 1.89; 95%CI (1.2 – 3.57)], lack of water supply to the household [AOR = 2.93; 95%CI (1.3 – 6.57)], and use of public toilet facility [AOR = 2.35; 95%CI (1.21 – 4.57)] were factors statistically significant with the outcome variables.

**Conclusion and Recommendation:** having upper respiratory tract infection in the past two weeks, not having water supply to the household, and using public toilet facilities had contributed to the risk of undernutrition. Strengthening screening of all children for undernutrition and improving water and sanitation services to the household are recommended.

## INTRODUCTION

One of the main causes of morbidity and mortality among children under the age of five, particularly in developing nations, is undernutrition. It includes underweight, which is typically caused by an inadequately balanced diet, chronic sickness during childhood, and stunted growth caused by persistent protein deficits. Nutrition is critical in the overall development of individuals and the nation at large. Despite its significance, malnutrition continues to be a serious health issue and is to account for one-third of all infants and child deaths, particularly in different countries. Malnutrition is also characterized as a condition that arises from consuming an unbalanced diet in which specific nutrients are absent, in excess (too high), or in the wrong proportion. Malnutrition is also defined as the ingestion of dietary nutrients either insufficiently or entirely (1).

According to UNICEF and WHO, third world countries are particularly affected by child malnutrition (2,3). Additionally, it was recognized that undernutrition contributes to child mortality (4). WHO reported that long-term malnutrition among children under the age of five results from poor dietary intake, which can negatively lead to dysfunction of the physical and mental health. Children who are severely malnourished are susceptible to impaired cognitive growth and development, which subsequently affects them as they age. The majority of the world’s African and Asian nations have malnourished children (3).

Of the 162 million children under the age of five who were stunted, 36% lived in Africa and 56% in Asia, according to the WHO. Acute respiratory infection (ARI), diarrhea, measles, and a few other infectious disorders are all connected with an increased risk of death when malnutrition is present. Malnutrition can occasionally seriously disrupt a child’s physical development and mental development, according to WHO (3).

Malnourished children are more likely to experience slowed physical and intellectual development, which reduces their adult productivity (5). Studies revealed that malnutrition among children under the age of five is linked to poor academic performance, increased absence from school, impaired intellectual accomplishment, delayed cognitive development, and increased illness morbidity and mortality (6).

Sub-Saharan Africa has high rates of child mortality due to a number of causes, including low caloric intake, high HIV/AIDS prevalence, political unrest, ineffective government policy execution, conflicts between different communities, etc. (1). For instance, 50% of children in Malawi, Burundi, and Madagascar are stunted due to insufficient nutritional intake or inadequate consumption of essential nutrients, according to UNICEF. Additionally, among 42 percent of children in Nigeria, malnutrition is the cause of 49 percent of school absences. Under-five children’s nutrition is impacted by a number of factors, including poverty, failure to exclusively breastfeed, maternal factors like poor nutrition during pregnancy, lack of appropriate weight gain, illnesses like diarrhea and acute respiratory infections, poor consumption of vitamin supplements or fortified foods, large family sizes, poor sanitation, lack of education and information about good or adequate nutrition, as well as food insecurity and safety (3).

Even if there has been a major improvement in the recent past, trends in children’s nutritional status suggest that Ethiopia still has a problem with child malnutrition (7). With an emphasis on children from low-income households who visit public health centers, this study was carried out to identify the determinant factors of child undernutrition among children aged 6 to 59 months in the Akaki Kaliti sub-city of Addis Abeba.

### Statement of the Problem

The utter elimination of extreme poverty and hunger is the goal of a continuous global effort. Malnutrition is still a significant public health issue, particularly in nations with little resources. Ninety percent of the world’s malnourished children lives in 36 developing countries (8,9).

One of the leading causes of morbidity and mortality among children worldwide is still malnutrition. It has been directly or indirectly accountable for 60% of the 10.9 million deaths of children under five that occur each year. These deaths, which are frequently linked to improper feeding habits, account for almost 23% of all infant deaths (10). Stunting, being underweight and wasting are all prevalent in Ethiopia at rates of 37%, 21%, and 7%, respectively. Fourteen percent of children in Addis Abeba are stunted, 5 percent are underweight, and 2 percent are wasted (7).

The nutritional status of children is impacted by a number of factors, including poverty, the absence of exclusive breastfeeding, maternal factors such as inadequate nutrition during pregnancy, inadequate weight gain, and inadequate vitamin supplement consumption, illness, environmental factors, socioeconomic/household factors, and environmental factors (1).

According to studies, the main risk factors for child undernutrition in low- and middle-income countries were household socioeconomic level and parental nutritional status. Child undernutrition was commonly linked to environmental factors, health habits, disease prevalence, and maternal reproductive care, with significant regional variation (7 - 9).

The weight of a child at birth, the mother’s age, her Body Mass Index (BMI), her marital status, and the identified regions with high incidence, such as Afar, Dire Dawa, Gambela, Harari, and Somali, were all linked to infant malnutrition in Ethiopia, according to a number of studies. Additionally, malnutrition was a problem for 48.5 percent of Ethiopian rural children (1). Children’s nutritional health in rural areas of the country was independently influenced by characteristics such age, gestational age, mother’s education level, and wealth status. The prevalence of stunting was 19.6%, wasting was 3.2 percent, and overweight/obesity was 11.4 percent among preschoolers in Addis Abeba. They also have limited access to high-quality foods and struggle with both under- and over-nutrition (10-16).

Between 2005 and 2019, there was a decrease in child undernutrition, according to trends in children’s nutritional status. Stunting has significantly decreased in frequency, from 51% in 2005 to 37% in 2019. Additionally, the prevalence of wasting dropped from 12 to 7 percent throughout the same time frame. Over these 14 years, there has been a constant decline in the percentage of underweight children, from 33% to 21% (7).

Malnutrition is a serious public health issue across the nation despite the fact that many different treatments against child malnutrition have been implemented in an effort to reduce those contributing factors. Therefore, it is crucial to determine the determinant variables that predispose the children to develop the condition in order to save children’s lives and the bottlenecks for reducing the prevalence of child undernutrition. Therefore, it is necessary to conduct research to identify the present prevalence and contributing factors of child malnutrition in Addis Abeba, Ethiopia, public hospitals.

## METHODS AND MATERIALS

### Study Area and Period

The study was conducted in Akaki Kaliti Sub city of Addis Ababa from April 26 to May 31, 2021. Addis Ababa is the capital city of Ethiopia and the African Union and is often called the “African Capital” due to its historical, diplomatic and political significance for the continent. It is also the largest city in the country by population. Akaki Kaliti Sub city is one of the 10 sub cities in Addis Ababa with a total population of 252,387 of which 18,071 are under five children. According to the Sub city health office report, the Sub city has one public hospital, one NGO hospital and 10 public health centers which are providing health care service to the population in the sub-city and the neighboring population of Oromia region.

### Study design

Facility-based case-control study was conducted.

### Population

#### Source Population

All children age 6 – 59 months with their mothers/care givers who are attending public health facility in Akaki Kaliti sub-city of Addis Ababa

### Study Population

#### Study population for cases

Children aged 6 – 59 months (cases) with their mothers/care givers who are attending public health facilities in Akaka Kaliti sub-city of Addis Ababa during the data collection period and who fulfilled the definition of case.

#### Study population for controls

Children aged 6 – 59 months (controls) with their mothers/care givers who are attending public health facilities in Akaka Kaliti sub-city of Addis Ababa during the data collection period and who have considered normal anthropometric measurement.

##### Definition of Cases

Cases were defined as any child between 6-59 months old presenting with any of the following World Health Organization anthropometric indicators for undernutrition. ***Stunted children*** were children with height/length –for-age z-score less than - 2 SD, ***wasted*** were children with weight – for-height z-score less than - 2 SD, and ***underweight*** were children with weight –for-age z-score less than - 2 SD and/or measurement of Mid Upper Arm Circumference (MUAC) <12cm.

##### Definition of Controls

Controls were defined as children between 6-59 months old considered having normal anthropometric indicators such as ***non-stunted*** children were children having height/length –for-age z-score greater than or equal to - 2 SD, ***not wasted*** children with weight –for-height z-score greater than or equal to - 2 SD, and ***not underweight*** children with weight –for-age z-score greater than or equal to - 2 SD and/or measurement of Mid Upper Arm Circumference (MUAC) greater than 12cm.

##### Inclusion Criteria

All children age 6 – 59 months with their index mothers/ care givers who attend the selected public health centers for services such as EPI, follow up, under 5 OPD and all other services were included

##### Exclusion Criteria

Children age 6 – 59 months who are acutely ill due to diseases other than malnutrition, with congenital anomaly, physical disability that can interfere with the anthropometric measurement, and children with edema were excluded

### Sample Size and Sampling Technique

#### Sample Size Determination

A double-population proportion formula using Epi-info version-7 was used to estimate the sample size required for the study. Sample size was calculated using four different exposure variables and the variable with largest sample size is taken. By considering the proportion of family size greater than five among controls was 57.2% (this variable was taken as the main exposure variable from previous study) (18), 95% CI, 80% power of the study, control to case ratio of 2:1 to detect an odds ratio of 2.28 (18). Accordingly, by adding 5% non-response rate, the total sample size using Fleiss w/cc method was **242** (**83 cases and 159 controls**).

#### Sampling Technique

Currently, there are two hospitals and 10 health centers found in Akaki Kaliti Sub-city of which all health centers were included in this study. According to the report of Akaki Kaliti sub-city health office, during the months of September to November 2020, a total of 4,131 children age 6 to 59 months were attended public health centers and screened for acute malnutrition of which 237 were diagnosed to have Acute malnutrition. Accordingly, the estimated sample size is allocated using probability proportional to size. Therefore, among children attending public health centers, the first case was selected by lottery method and then enrolled using consecutive sampling technique until the estimated sample size was achieved (See table 2)

**Table 1:**
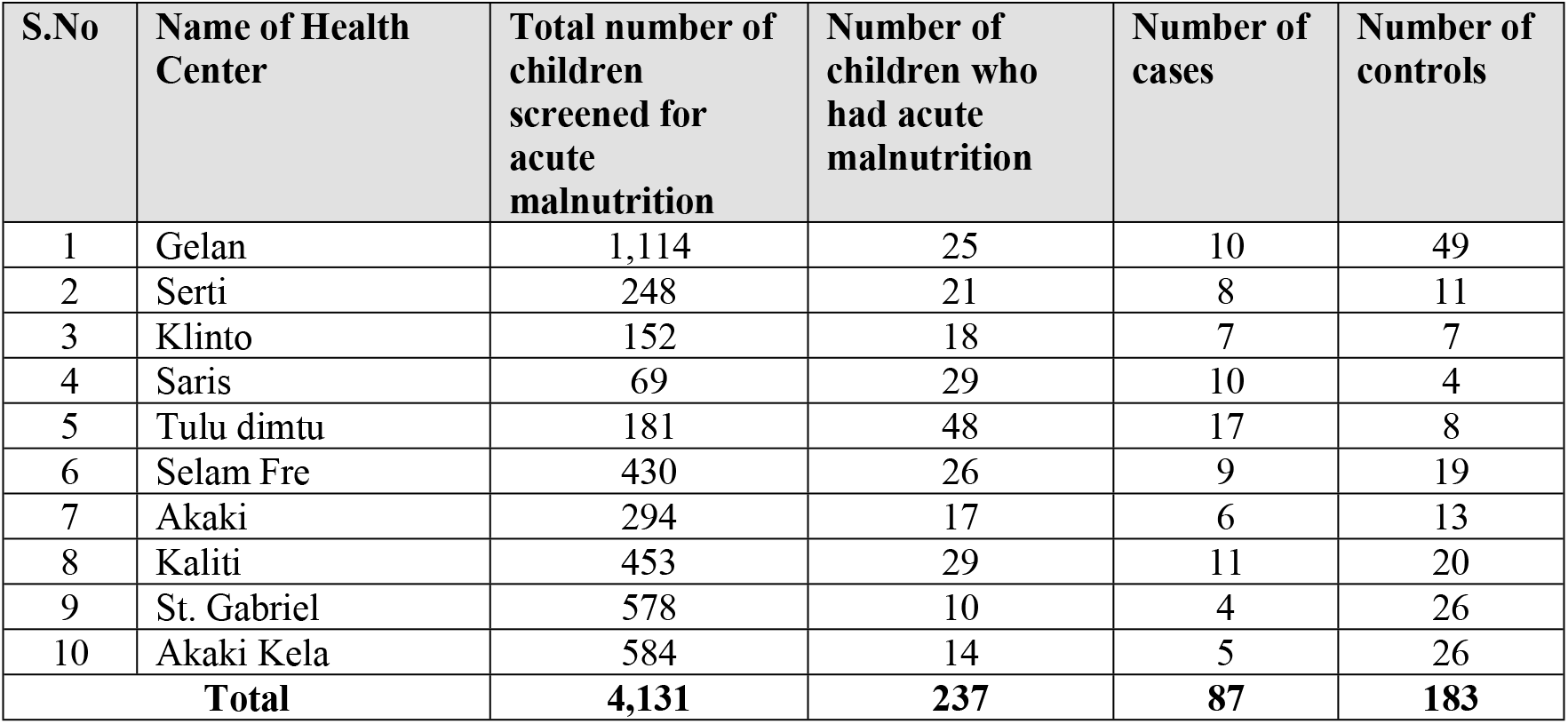
Sample Size distribution using probability proportional to the size

**Table 2:** Socio-demographic Characteristics of Respondents (n=242)

| **Variables** | **Total**<br>**#(%)** | **Cases**<br>**# (%)** | **Controls**<br>**#(%)** | ***X<sup>2</sup> test*** | **P-value** |
| --- | --- | --- | --- | --- | --- |
| **Age of the mother /care giver in years** |  |  |  |  |  |
| 15-19 Year | 3 (1.2) | 2 (2.5) | 1 (0.6) |  |  |
| 20-24 Year | 36 (14.9) | 9 (11.2) | 27 (16.7) |  |  |
| 25-29 Year | 92 (38.0) | 31 (38.8) | 61 (37.7) | 3.2235 | 0.780 |
| 30-34 Year | 73 (30.2) | 26 (32.5) | 47 (29.0) |  |  |
| 35-39 Year | 27 (11.2) | 8 (10.0) | 19 (11.7) |  |  |
| 40-44 Year | 9 (3.7) | 3 (3.8) | 6 (3.7) |  |  |
| 45 & above | 2 (0.8) | 1 (1.2) | 1 (0.6) |  |  |
| **Educational status of the mother/care giver** |  |  |  |  |  |
| No formal education | 43 (17.8) | 23 (28.7) | 20 (12.3) |  |  |
| Primary education | 87 (35.9) | 22 (27.5) | 65 (40.1) | 12.4488 | 0.006 |
| Secondary education | 63 (26.0) | 23 (28.7) | 40 (24.7) |  |  |
| College and above | 49 (20.2) | 12 (15.0) | 37 (22.8) |  |  |
| **Current marital status** |  |  |  |  |  |
| Married | 193 (79.8) | 64 (80.0) | 129 (79.6) |  |  |
| Single | 33 (13.6) | 13 (16.2) | 20 (12.3) | 2.4345 | 0.487 |
| Divorce | 3 (1.2) | 1 (1.2) | 2 (1.2) |  |  |
| Widowed | 13 (5.4) | 2 (2.5) | 11 (6.8) |  |  |
| **Religion** |  |  |  |  |  |
| Orthodox | 146 (60.3) | 53 (66.2) | 92 (56.8) |  |  |
| Muslim | 52 (21.5) | 16 (20.0) | 36 (22.2) | 3.3785 | 0.337 |
| Catholic | 6 (2.5) | 1 (1.2) | 5 (3.1) |  |  |
| Protestant | 38 (15.7) | 9 (11.2) | 29 (17.9) |  |  |
| **Current occupation of the mother/care giver** |  |  |  |  |  |
| Housewife | 122 (50.4) | 38 (47.5) | 82 (50.6) |  |  |
| Civil servant | 45 (18.6) | 12 (15.0) | 33 (20.4) | 5.9644 | 0.113 |
| Business woman | 44 (18.2) | 12 (15.0) | 31 (19.1) |  |  |
| Daily labor | 31 (12.8) | 14 (17.5) | 15 (9.3) |  |  |
| **Sex of child** |  |  |  |  |  |
| Male | 145 (59.9) | 50 (62.5) | 95 (58.6) | 0.3319 | 0.565 |
| Female | 97 (40.1) | 30 (37.5) | 67 (41.4) |  |  |
| **Age of child** |  |  |  |  |  |
| 6-23 month | 127 (52.5) | 36 (45) | 91 (56.2) | 2.6807 | 0.102 |
| 24 -59 month | 115 (47.5) | 44 (55) | 71 (43.8) |  |  |

### Study Variables

#### Dependent Variable

- Child undernutrition status (Cases/Controls)

#### Independent Variables

- Sociodemographic and Socio-economic factors: includes sex, age of mother in years, age of the child in months, family size, current marital status, sex of the household head, and whether the mother was involved in an income-earning activity.
- Child related factors: age, sex, disease status
- Maternal factors: breast feeding and positioning, education, age, house hold income
- Socioeconomic/household factors: family size, household income, parent education, lack of access to food
- Environmental factors: residence, water supply

### Data Collection Procedure

#### Data Collection Instrument

The questionnaire for this study was adapted from the relevant literatures based on the study objective. The questionnaire includes socioeconomic and demographic factors, child feeding and caring practices, maternal health factors, environmental health related characteristics, and anthropometrics measurements. The participant’s socio-demographic characteristics includes sex (male or female), age of mother in years, age of the child in months, family size, current marital status (married/living together, divorced/widowed/separated), sex of the household head (female/male), and whether the mother was involved in an income-earning activity (yes/no).

Socioeconomic characteristics were maternal education, household wealth, and household food security. The maternal education was about the highest level of schooling completed by the mother at the time of the survey. The level of education was then have grouped into five categories: never attended/not finished first grade, grade 1–4, grade 5–8, grade 9–12, and college-educated. The household wealth was ownership of house, type of housing unit, access to separate toilet facility, refrigerator, bed, and stove.

#### Anthropometric measurements of the children

Length/height was measured without shoes, socks, hair/head scurf, and ornaments and positioning the subject at the Frankfurt plane by using wooden board inserted with a tape calibrated to read the nearest 0.1cm. Length of the infants was measured in a recumbent (lying) position using a horizontal wooden length board and movable headpiece. Height was measured for children older than two years of age in standing position into the nearest 0.1 cm using a vertical wooden height board by placing the child on the measuring board, and child standing upright in the middle of board and head held erect such that the external auditory meatus and the lower boarder of the eye were in one horizontal plane (Frankfurt plane). Anthropometric measurement was taken twice and a difference of 0.1 cm in length was accepted as normal.

#### Data Collection Technique

One diploma nurse with prior data collection experience was recruited for each health center and two BSc public health officer were recruited as a supervisor and each of them supervised five health centers. Diagnosis of acute malnutrition was undertaken using WFH<80% and/or their MUAC<12.5 cm (MAM) and WFH<70% and/or MUAC<11.5 cm (SAM) with or without bilateral pitting edema to identify eligible and non-eligible subjects as well as to classify those eligible children as cases and controls by experienced public health officer who was assigned at each health center during the study period.

After screening of cases and controls, data was collected using an interviewer-administered structured questionnaire. After informed verbal consent was obtained, data was collected from index mothers or care givers of children 6–59 months of age.

#### Data Quality Management (Data Quality Assurance and Data Quality Control)

Data quality assurance was made through training of data collators, questionnaire pretesting and continuous supervision at the time of data collection. A day long training was given for data collectors and supervisor before the actual data collection period. The training was focus on anthropometric measurement techniques, study objectives, introducing the data collection tool, how to approach study participants, wise use of time and each measurement should be standardized by taking twice and validated by calculating technical error of measurement (TEM). The weight of the children was measured to the nearest 0.1 kg using the United Nations International Children’s Emergency Fund (UNICEF) electronic scale. MUAC was taken using the WHO-recommended MUAC tape and procedure, and recumbent length and height was measured to the nearest 0.1 cm using UNICEF’s recommended model wooden board, as per the WHO protocol. Daily supervision was held at study settings by supervisor and principal investigator. The questionnaire was pretested on 5% of the sample in separate health center and possible modification was made before the actual data collection as needed. The data collection tool was checked each day during the actual data collection time for completeness and consistency by supervisor and principal investigator. The code was given in completed questionnaire. The data on coded questionnaires was entered into Epi-data version 3.1 and WHO Anthro software (WHO, Geneva, Switzerland) was used to convert weight, height and age data into Z-scores using the 2006 WHO Growth Standards by the principal investigator using a double entry method(21).

#### Data Processing and Analysis

Data was cleaned, assessed for missing data and entered using Epi-data version 3.1 and analyzed using SPSS for windows version 21. Continuous explanatory variables were categorized. Descriptive analysis was undertaken and the result was presented by tables and graphs. Bivariate analyses were performed to assess the association of each independent variable with the outcome variable and to identify candidate variables for multivariable analysis. Candidate variables with p-value less than 0.25 in the bivariate analysis was entered to multivariable logistic regression model to identify independent predictors of child malnutrition and the magnitude of association between the different variables in relation to the outcome variable was measured by adjusted odds ratio (AOR) with 95% confidence interval (CI). A P-value less than 0.05 at 95% CI as cut of point was used to declare the observed association is statistically significant.

#### Operational Definitions

***Stunted:*** a child with a HAZ less than −2 SD

***Wasted:*** a child with WHZ less than −2 SD from the reference population

***Underweight***: a child with WHZ less than – 2 SD from the reference population ***Overweight/Obese:*** a child with WHZ less than +2 SD from the reference population **Ethical**

#### Considerations

The ethical approval and clearance for the study was obtained from Institutional Review Board (IRB), Menelik-II College of health science, Kotebe Metropolitan University. An official letter of permission was obtained from each health facility included in the study. Informed verbal consent was obtained from the study participants after explaining the purpose of the study. To assure confidentiality, name of the participant was not written on the questionnaire.

#### Dissemination Plan

Results of this study will be presented to the Menelik-II College of health science, Kotebe Metropolitan University. The findings will be delivered through hard and soft copy to health centers included in the study, Addis Ababa city administration health bureau, Federal Ministry of Health, and other concerned stakeholders. Repeated discussion and policy brief will be considered as needed. Efforts will be made to present the results on international and national professional conferences, and publication on peer reviewed scientific journal will also have considered.

## RESULTS

### Socio-demographic Characteristics of Subjects

A total of 242 subjects (83 cases and 159 controls) were included in the study giving the overall response rate of 94.2%. The mean age (±SD) of the mother/care givers of cases was 29.59 ± 5.588 years and 29.46 ± 5.0 for controls. The mean age (±SD) of cases was 24.17 ± 14.429 months and 24.74 ± 14.67 months for control. Of the total, majorities, 64(80%) of mothers/care givers of cases as compared to 129(79.6%) of mothers/care givers of controls were married. Regarding religion, 53(63.2%) of cases and 92(56.8%) of controls were orthodox believers. Regarding, educational status of the mother/care giver, 23(28.7%) of cases and 20(12.3%) of controls did not attend formal education. About half, 38(47.5%) of mothers/care givers of cases and 82(50.6%) of controls were housewives in their current occupation (See table 3)

**Table 3:** Recent childhood illness related factors of undernutrition among children age 6 to 59 months in Akaki Kaliti Sub-city, Addis Ababa, Ethiopia, 2021(n=242)

| Variables | Total # (%) | Cases # (%) | Controls # (%) | $X^2$ test | P-value |
| --- | --- | --- | --- | --- | --- |
| **URTI in the past two weeks** |  |  |  |  |  |
| No | 163 (67.4) | 47 (58.8) | 116 (71.6) | 4.0248 | 0.045 |
| Yes | 79 (32.6) | 33 (41.2) | 46 (28.4) |  |  |
| **Diarrhea in the past two weeks** |  |  |  |  |  |
| No | 199 (82.2) | 62 (77.5) | 137 (85.1) | 1.8310 | 0.176 |
| Yes | 43 (17.8) | 18 (22.5) | 24 (14.9) |  |  |
| **Vaccination** |  |  |  |  |  |
| No | 12 (5.0) | 3 (3.8) | 9 (5.6) | 0.3705 | 0.543 |
| Yes | 230 (95.0) | 77 (96.2) | 153 (94.4) |  |  |

Regarding demographic characteristics of the child, 50(62.5%) of cases and 95(58.6%) controls were male in sex, and 36(45%) of cases and 91(56.2%) controls were in the age range of 6 to 23 months.

### Recent Childhood Illness

Of the total study subjects, 33(41.2%) of cases and 46(28.4%) of controls children had upper respiratory infection in the last two weeks; and 18(22.5%) of cases and 24(14.9%) of control children had diarrhea in the past two weeks. Regarding vaccination status, nearly all, 77(96.2%) of cases and 153(94.4%) of controls were fully vaccinated (see table 4)

**Table 4:**
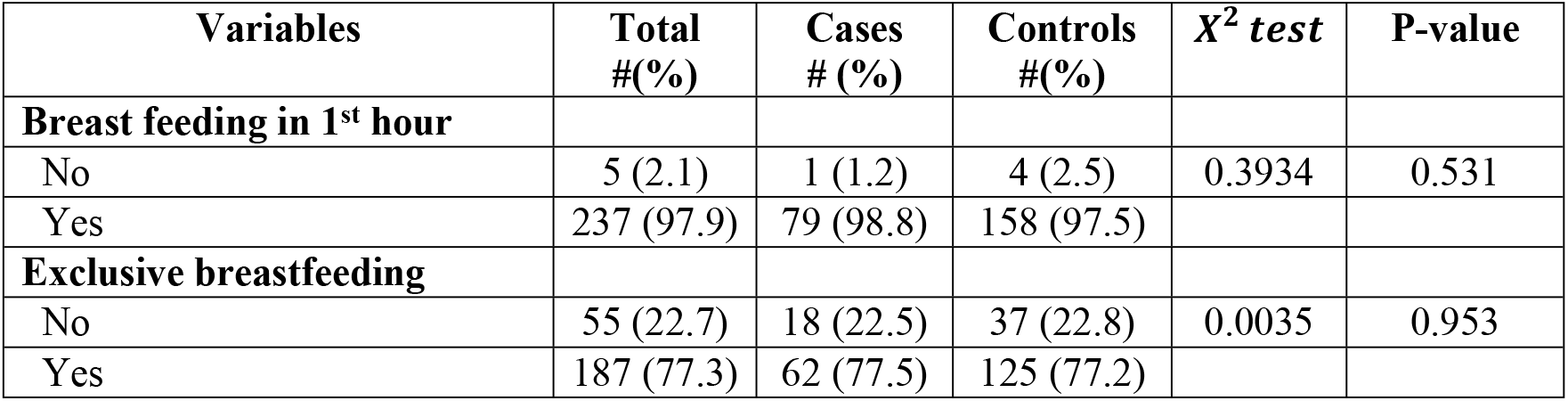
Breast feeding related factors of undernutrition among children age 6 to 59 months in Akaki Kaliti Sub-city, Addis Ababa, Ethiopia, 2021(n=242)

| Variables | Total # (%) | Cases # (%) | Controls # (%) | $X^2$ test | P-value |
| --- | --- | --- | --- | --- | --- |
| **Breast feeding in 1<sup>st</sup> hour** |  |  |  |  |  |
| No | 5 (2.1) | 1 (1.2) | 4 (2.5) | 0.3934 | 0.531 |
| Yes | 237 (97.9) | 79 (98.8) | 158 (97.5) |  |  |
| **Exclusive breastfeeding** |  |  |  |  |  |
| No | 55 (22.7) | 18 (22.5) | 37 (22.8) | 0.0035 | 0.953 |
| Yes | 187 (77.3) | 62 (77.5) | 125 (77.2) |  |  |

### Breast feeding

Regarding breast feeding of a children, almost all, 79 (98.8%) of mothers/care givers of cases and 158(97.5%) of controls breast fed their children with in the first hour after birth. Nearly two-third, 62(77.5%) of mothers/care givers of cases and 125 (77.2%) of control children had exclusive breast feeding (see table 5)

**Table 5:**
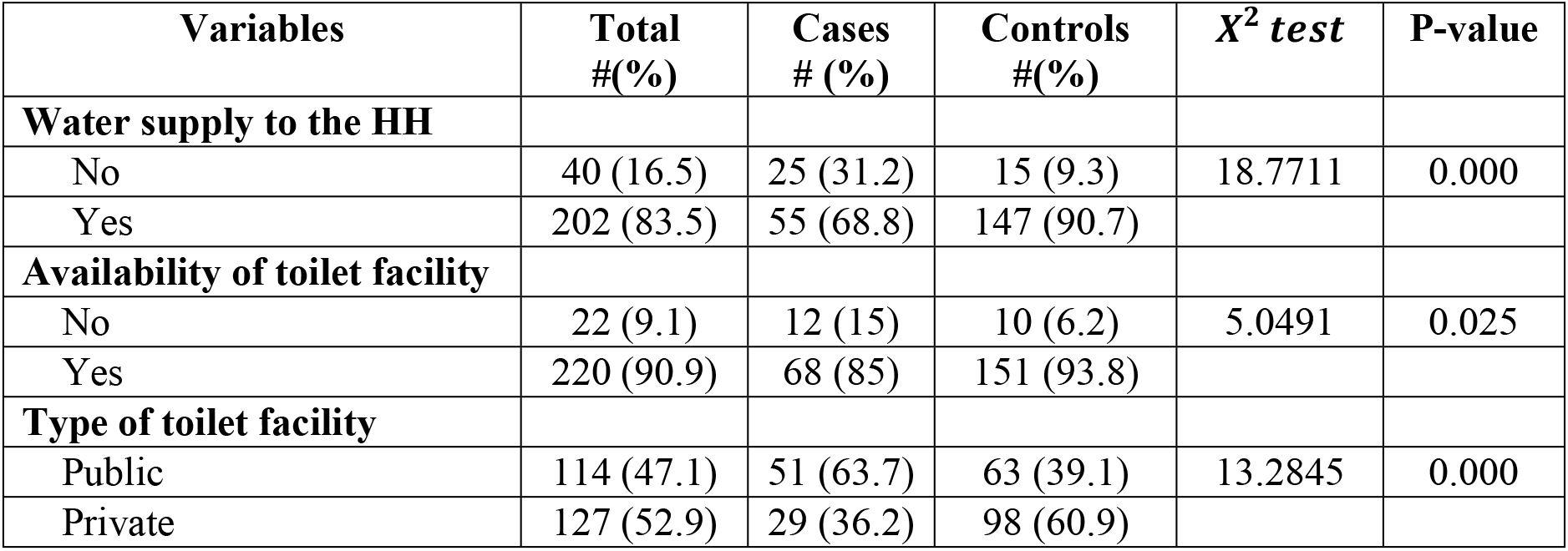
Water and sanitation related factors of undernutrition among children age 6 to 59 months in Akaki Kaliti Sub-city, Addis Ababa, Ethiopia, 2021(n=242)

### Water and Sanitation

Regarding environmental factors of child undernutrition, about 55(68.8%) of cases and 147(90.7%) of controls have water supply to their household, 68(85%) of cases and 151(93.8%) of control subjects have toilet facility, and of which, 29 (36.2%) of cases and 98(60.9%) of control subjects have private toilet facility (see table 6)

**Table 6:** Types of undernutrition among children age 6 to 59 months in Akaki Kaliti Sub-city, Addis Ababa, Ethiopia, 2021(n=242)

| Level of malnutrition | Age of child |  |  | *X<sup>2</sup> test* | P-value |
| --- | --- | --- | --- | --- | --- |
|  | 6 - 23 Months | 24 - 59 Months | Total |  |  |
| **Underweight (WAZ)** |  |  |  |  |  |
| Moderate | 17(47.2) | 15(34.1) | 32(40.0) | 1.4226 | 0.233 |
| Sever | 19(52.8) | 29(65.9) | 48(60.0) |  |  |
| **Wasting (WHZ)** |  |  |  |  |  |
| Normal | 14(38.9) | 16(36.4) | 30(37.5) |  |  |
| Moderate | 8 (22.2) | 11(25.0) | 19(23.8) | 0.0983 | 0.952 |
| Sever | 14(38.9) | 17(38.6) | 31(38.8) |  |  |
| **Stunting (HAZ)** |  |  |  |  |  |
| Normal | 10(27.8) | 13(29.6) | 23(28.8) |  |  |
| Moderate | 10(27.8) | 5 (11.4) | 15(18.8) | 3.6757 | 0.159 |
| Sever | 16(44.4) | 26(59.1) | 42(52.5) |  |  |

NB: Normal = Z-score less than -2 SD, Moderate = Z-score ( $\leq -3$ to $\geq -2$ ) SD, Sever = Z-score $\leq -3$

### Measurement of Under-nutrition Status

#### Underweight

regarding undernutrition status as shown in table 7 below, this study identified that, among all underweight children, moderate underweight were 17 (47.2%) in the age range of 6 – 23 months whereas 15 (34.1%) were between 24 – 59 months. Likewise, 19 (52.8%) and 29(65.9%) of children with severe underweight were in the age range of 6 – 23 months and 24 – 59 months, respectively.

**Table 7:** Predictors of undernutrition among children age 6 to 59 months in Akaki Kaliti Sub-city, Addis Ababa, Ethiopia, 2021(n=242)

| **Variables** | **Cases**<br>**# (%)** | **Controls**<br>**# (%)** | **COR (95%CI)** | **P-value** | **AOR (95%CI)** | **P-value** |
| --- | --- | --- | --- | --- | --- | --- |
| **Age of mother / care giver** |  |  |  |  |  |  |
| 15 -29 years | 44(53) | 87(54.7) | ref |  | ref |  |
| Above 30 years | 39(47) | 72(45.3) | 1.1 (0.64,1.89) | 0.720 | 1.31 (0.7,2.46) | 0.392 |
| **Maternal education** |  |  |  |  |  |  |
| No formal education | 23 (28.7) | 20 (12.3) | 3.55 (1.46,8.59) | 0.005* | 2.23 (0.62,8.0) | 0.219 |
| Primary education | 22 (27.5) | 65 (40.1) | 1.04 (0.46,2.35) | 0.918 | 0.92 (0.30,2.86) | 0.886 |
| Secondary education | 23 (28.7) | 40 (24.7) | 1.77 (0.77,4.06) | 0.176 | 1.76 (0.61,5.11) | 0.296 |
| College and above | 12 (15.0) | 37 (22.8) | ref |  | ref |  |
| **Maternal Occupation** |  |  |  |  |  |  |
| Housewife | 38 (47.5) | 82 (50.6) | ref |  | ref |  |
| Civil Servant | 12 (15.0) | 33 (20.4) | 0.77 (0.36,1.66) | 0.510 | 1.44 (0.48,4.33) | 0.516 |
| Business woman | 12 (15.0) | 31 (19.1) | 0.89 (0.42,1.89) | 0.767 | 1.24 (0.51,3.04) | 0.632 |
| Daily laborer | 14 (17.5) | 15 (9.3) | 2.27 (1.02,5.06) | 0.045* | 2.38 (0.94,6.00) | 0.067 |
| **Sex of the child** |  |  |  |  |  |  |
| Male | 50 (62.5) | 95 (58.6) | 1.18 (0.68,2.04) | 0.565 | 0.9 (0.49,1.66) | 0.741 |
| Female | 30 (37.5) | 67 (41.4) | ref |  | ref |  |
| **Age of child** |  |  |  |  |  |  |
| 6- 23 months | 36 (45) | 91 (56.2) | ref |  | ref |  |
| 24 - 59 months | 44 (55) | 71 (43.8) | 1.57 (0.91,2.68) | 0.103 | 1.53 (0.84,2.8) | 0.162 |
| **Breast feeding in the 1<sup>st</sup> hr** |  |  |  |  |  |  |
| No | 1 (1.2) | 4 (2.5) | 0.5 (0.05,4.55) | 0.538 | 0.48 (0.05,4.74) | 0.530 |
| Yes | 79 (98.8) | 158 (97.5) | ref |  | ref |  |
| **Exclusive breast feeding** |  |  |  |  |  |  |
| No | 18 (22.5) | 37 (22.8) | 0.98 (0.52,1.86) | 0.953 | 0.79 (0.38,1.62) | 0.517 |
| Yes | 62 (77.5) | 125 (77.2) | ref |  | ref |  |
| **URTI in the past two weeks** |  |  |  |  |  |  |
| Yes | 47 (58.8) | 116 (71.6) | 1.77 (1.01,3.1) | 0.046* | **1.89 (1.2,3.57)** | **0.050\*** |
| No | 33 (41.2) | 46 (28.4) | ref |  | ref |  |
| **Diarrhea in the past two weeks** |  |  |  |  |  |  |
| Yes | 18(22.5) | 24(14.9) | 0.97(0.67,1.43) | 0.15 | 0.95(0.55,1.63) | 0.144 |
| No | 62(77.5) | 137(85.1) | ref |  | ref |  |
| **Water supply to the HH** |  |  |  |  |  |  |
| Yes | 55 (68.8) | 147 (90.7) | ref |  | ref |  |
| No | 25 (31.2) | 15 (9.3) | 4.45 (2.19,9.07) | < 0.001* | **2.93 (1.3,6.57)** | **0.009\*** |
| **Availability of toilet facility** |  |  |  |  |  |  |
| Yes | 68 (85) | 151 (93.8) | ref |  | ref |  |
| No | 12 (15) | 10 (6.2) | 0.37(0.15,0.91) | 0.02* | 0.37(0.13,1.04) | 0.07 |
| **Type of toilet facility** |  |  |  |  |  |  |
| Public | 51 (63.7) | 63 (39.1) | 2.76 (1.59,4.81) | < 0.001* | **2.35 (1.21,4.57)** | **0.011\*** |
| Private | 29 (36.2) | 98 (60.9) | ref |  | ref |  |

\*: Significant at $\alpha=0.005$ ; COR: Crude Odds Ratio; AOR: Adjusted Odds Ratio; CI: Confidence Interval

#### Stunting

similarly regarding stunting, 8 (22.2%) and 11(25%) of children with moderate stunting were in the age range of 6 – 23 months and 24 – 59 months, respectively. Also, 14 (38.9%) and 17 (38.6%) of children with severe stunting were in the age range of 6 – 23 months and 24 – 59 months, respectively (table 7).

#### Wasting

regarding wasting status of the children, about 19 (23.8%) of children were found to have moderate wasting whereas 31 (38.8%) have severe wasting (table 7)

#### Predictors of Child Undernutrition

In this study, bivariate analysis identified a total five candidate variables for multivariable analysis. After controlling for potential confounding as shown in table 7, this study identified three independent predictors of child undernutrition.

This study revealed that upper respiratory tract infection in the past two weeks showed statistically significant association with undernutrition. The odds of undernutrition among children who had upper respiratory tract infection in the past two weeks was nearly two times higher as compared to those children who did not have upper respiratory tract infection [AOR = 1.89; 95%CI (1.2 – 3.57), p-value = 0.05].

This study also indicated that lack of water supply to the household showed statistically significant association with child undernutrition. The odds of undernutrition among children who didn’t have water supply to their household were approximately three times higher as compared to those who have adequate water supply to their household [AOR = 2.93; 95%CI (1.3 – 6.57), p-value = 0.009]

Consequently, this study disclosed that type of toilet facility was significantly associated with child undernutrition. Specifically, the odds of undernutrition children using public toilet facility was 2.3 times higher as compared to those using private toilet facility [AOR = 2.35; 95%CI (1.21 – 4.57), p-value= 0.01].

## DISCUSSION

This study revealed upper respiratory tract infection in the past two weeks, lack of water supply to the household, and using public toilet facility are independent positive predictors of child undernutrition.

According to this study, the odds undernutrition among children who had upper respiratory tract infection two weeks preceding the study was about two times higher as compared to those children who did not have upper respiratory tract infection. This finding is consistent with the study finding conducted in Gondar city, Northwest Ethiopia (30). The implication of this finding might be that during illness the metabolic rate could increase due to the disease process such as fever, which might predispose the child to the development of undernutrition (33).

This study also identified that water supply to the house hold is a determinant factor for child undernutrition. Specifically, the odds of undernutrition among children who don’t have water supply to their household were approximately three times higher as compared to those who have adequate water supply to their household. This finding is supported by the finding from the study conducted in Oromia region, West Ethiopia (33). This might imply that inadequate water supply imposes a problem of sanitation which predisposes children to acquire different types of soil transmitted helminths and diarrheal diseases thus increase the likelihood of undernutrition (34)

According to this study, the odds of undernutrition among children who are using public toilet facility were about 2.3 times higher as compared to their counter parts. This finding is in line with other previous studies conducted in Narok city, Kenya and Gambella town, Southwest Ethiopia (31, 32). The possible implication of this finding might be that using public toilet facility predispose the family members for cross-contamination which in turn increases the likelihood of undernutrition in the children (31,32)

### Strength and Limitation of the Study

This study has its own strengths and limitations. Regarding strengths, this study utilized primary data and applied a stronger study design which is reasonably appropriate to identify determinant factor.

Concerning limitations of this study, the first potential limitation is of recall bias. Because of the nature of the study design employed, exposure assessment was done retrospectively, so that there was possibility of recall bias. In addition, this study was limited public health facilities.

## CONCLUSION AND RECOMMENDATION

### Conclusion

In conclusion, this study has found child health related and environmental factors had contributed to the risk of undernutrition. Upper respiratory tract infection in the past two weeks, lack of water supply to the household, and using public toilet facility were identified as independent positive predictors of child undernutrition.

On the other hand, age of the mother/care giver, family size, current marital status, age of the child, sex of the child breast feeding in the first hour after birth, exclusive breast feeding, educational status of the mother/care giver, residence and availability of toilet facility to the household were not identified as determinants of child undernutrition.

### Recommendation

#### To Akaki Kaliti Health Office / Health Bureau of City Administration of Addis Ababa

- Strengthen screening of all children age 6 to 59 months for undernutrition in all health facilities.
- Strengthen health education session both in facility and community level about the consequences of child undernutrition and prevention methods.

#### To researchers

- Since the scope of this study was limited to public health facilities further large-scale research which encompasses private and NGO facilities is recommended.

### Data Availability

The data used to support the findings of this study are available from the corresponding author upon request.

## Authors’ Contributions

All authors were equally contributed in design, coordination, reviewed the article for the study, and report writing. All authors have been contributed for works like; analyzing the data, drafting and writing the final manuscript. All authors have read and approved the final manuscript.

## Acknowledgments

We want to extend our appreciation to Institutional Review Board (IRB), Menelik-II College of health science, Kotebe Metropolitan University. We would like to express our deep appreciation for research participants, and data collectors for their cooperation.

## ACRONYMS/ABBREVIATIONS

AIDS: Acquired Immune Deficiency Syndrome
ARI: Acute Respiratory Infection
BMI: Body Mass Index
EDHS: Ethiopia Demographic Health Survey
FANTA: Food and Nutrition Technical Assistance
HAZ: Height-for-Age Z-score
HIV: Human Immunodeficiency Virus
SD: Standard Deviation
TB: Tuberculosis
UNICEF: United Nations Children’s Fund
WHO: World Health Organization
WHZ: Weight-for-Height Z-score
HC: Health Center

